# Presenting features of COVID-19 in older people: relationships with frailty, inflammation and mortality

**DOI:** 10.1101/2020.06.07.20120527

**Authors:** Paul Knopp, Amy Miles, Thomas E Webb, Benjamin C Mcloughlin, Imran Mannan, Nadia Raja, Bettina Wan, Daniel Davis

## Abstract

**Purpose:** To describe the clinical features of COVID-19 in older adults, and relate these to outcomes.

**Methods:** Cohort study of 217 individuals (≥70 years) hospitalised with COVID-19, followed up for allcause mortality. Secondary outcomes included cognitive and physical function at discharge. C-reactive protein and neutrophil: lymphocyte ratio were used as measures of immune activity.

**Results:** Cardinal COVID-19 symptoms (fever, dyspnoea, cough) were common but not universal. Inflammation on hospitalisation was lower in frail older adults. Fever, dyspnoea, delirium and inflammation were associated with mortality. Delirium at presentation was an independent risk factor for cognitive decline at discharge.

**Conclusions:** COVID-19 may present without cardinal symptoms as well as implicate a possible role for age-related changes in immunity in mediating the relationship between frailty and mortality.

**Key summary points:** *Aim:* To characterise symptoms, key findings and clinical outcomes in older adults with COVID-19

*Findings:* 12% of older individuals did not present with classical COVID-19 symptoms, though fever, dyspnoea, delirium and raised inflammation were associated with higher mortality. Compared with fitter older individuals, immune activity was lower in frailer patients.

*Message:* COVID-19 may present without cardinal symptoms as well as implicate a possible role for age-related changes in immunity in mediating the relationship between frailty and mortality.

## Introduction

As healthcare systems around the world start to respond to SARS-CoV-2, a major consideration is the apparent age-related heterogeneity in presentation, treatment responsiveness and clinical outcomes [1]. COVID-19 in older people, *prima facie*, may present in the absence of classical symptoms, progress more rapidly to severe disease, have poorer intensive care outcomes, longer inpatient stay and higher mortality [2, 3]. Nonetheless, questions remain as to which presenting features have greatest impact on these outcomes in older people. Recognising this may lead to better clinical care, as well as forming the basis for new services for older people after COVID-19.

Given this urgent need to understand COVID-19 in older people, we set out to describe the clinical, laboratory and radiological features in a series of older individuals hospitalised with COVID-19 in a large urban hospital.

## Methods

### Study design and participants

We undertook a prospective cohort study of patients aged ≥70 years admitted to University College Hospital, a large central London hospital, diagnosed with COVID-19 up until 23rd April 2020. Patients were included if they tested positive for SARS-CoV-2 on reverse-transcriptase polymerase chain reaction from a combined oropharyngeal and nasal swab. We also included swab-negative participants with a clinical diagnosis of COVID-19 on review of clinical, laboratory and radiological findings by a specialist infectious diseases team.

### Outcome

Primary outcome was all-cause mortality recorded during admission or if occurring after discharge updated from NHS Spine, a collection of local and national demographic databases. Vital status was followed up until 13th May 2020. Secondary outcomes included any decreased cognitive or physical function at discharge.

### Clinical measures

We recorded demographic data on age, sex and ethnicity. Presenting features included fever, cough, dyspnoea, and gastrointestinal symptoms, along with any geriatric syndromes: delirium, reduced mobility, and falls. The Clinical Frailty Scale (CFS) was used to quantify frailty, with scores assigned by specialist geriatricians. We measured C-reactive protein (CRP) and neutrophil: lymphocyte ratios as indicators of immune activity and noted the presence of any radiological abnormalities reported by specialist radiologists.

### Statistical analyses

We regarded presenting features as being present or absent. We examined distributions of CRP and neutrophil: lymphocyte ratios at graded levels of frailty (CFS 1 to 3; CFS 4 to 6; CFS 7 to 9), with differences in median values assessed using the Kruskal-Wallis test. We estimated associations between presenting clinical, laboratory or radiological features with mortality in a series of univariable and multivariable Cox proportional hazards models. To estimate associations with increased rehabilitation needs (cognitive and/or physical), we used logistic regression. Post-estimation procedures included Schoenfeld residuals and Hosmer-Lemeshow tests for heteroskedasticity. Stata 14.1 (StataCorp, Texas, USA) was used for all analyses.

## Results

We identified 217 individuals aged ≥70 years hospitalised with COVID-19. Mean age of patients was 80 years (range 70 to 99 years), 62% were male and a range of pre-morbid frailty was identified (Table 1). The majority of patients had no formal package of community care (n=154, 71%) and 8 patients (4%) were admitted from residential care homes. 72 individuals (33%) were living with definite or probable dementia (Table 1).

**Table 1.**
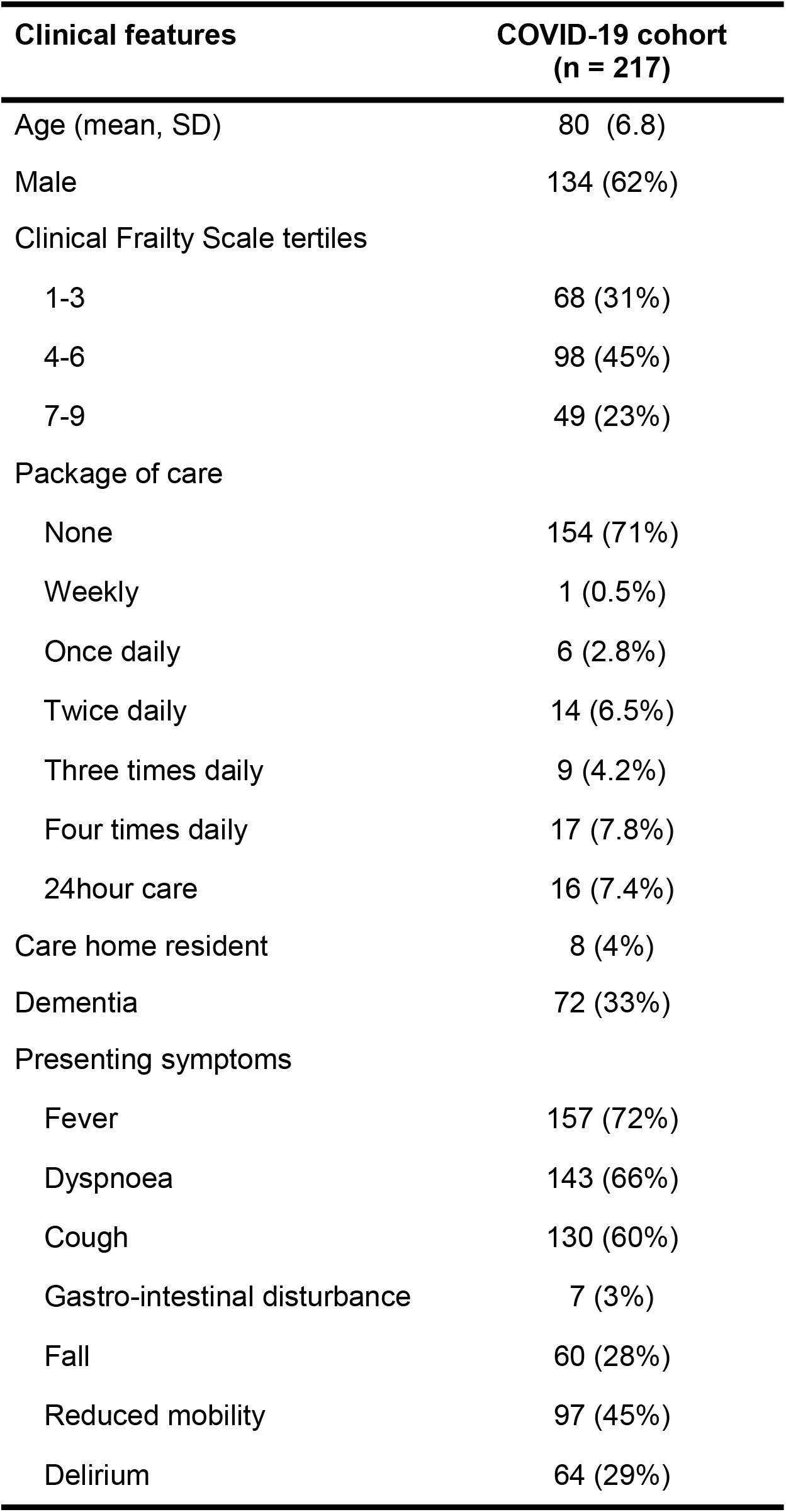
Characteristics and presenting symptoms of older adults (≥70 years) with COVID-19 admitted to hospital

| **Clinical features** | **COVID-19 cohort**<br>**(n = 217)** |
| --- | --- |
| Age (mean, SD) | 80 (6.8) |
| Male | 134 (62%) |
| Clinical Frailty Scale tertiles |  |
| 1-3 | 68 (31%) |
| 4-6 | 98 (45%) |
| 7-9 | 49 (23%) |
| Package of care |  |
| None | 154 (71%) |
| Weekly | 1 (0.5%) |
| Once daily | 6 (2.8%) |
| Twice daily | 14 (6.5%) |
| Three times daily | 9 (4.2%) |
| Four times daily | 17 (7.8%) |
| 24hour care | 16 (7.4%) |
| Care home resident | 8 (4%) |
| Dementia | 72 (33%) |
| Presenting symptoms |  |
| Fever | 157 (72%) |
| Dyspnoea | 143 (66%) |
| Cough | 130 (60%) |
| Gastro-intestinal disturbance | 7 (3%) |
| Fall | 60 (28%) |
| Reduced mobility | 97 (45%) |
| Delirium | 64 (29%) |

On admission, symptoms of fever, dyspnoea and cough were common (n=157, 72%; n=143, 66%; n=130, 60% respectively) (Table 1). Symptoms of gastro-intestinal disturbance were less frequent (n=7, 3%). Some individuals were admitted without any of these cardinal COVID-19 symptoms (n=25, 12%), instead presenting with one or more frailty syndrome (reduced mobility, falls or delirium). Figure 1 details the combinations of presenting symptoms observed in our sample.

**Fig. 1.**
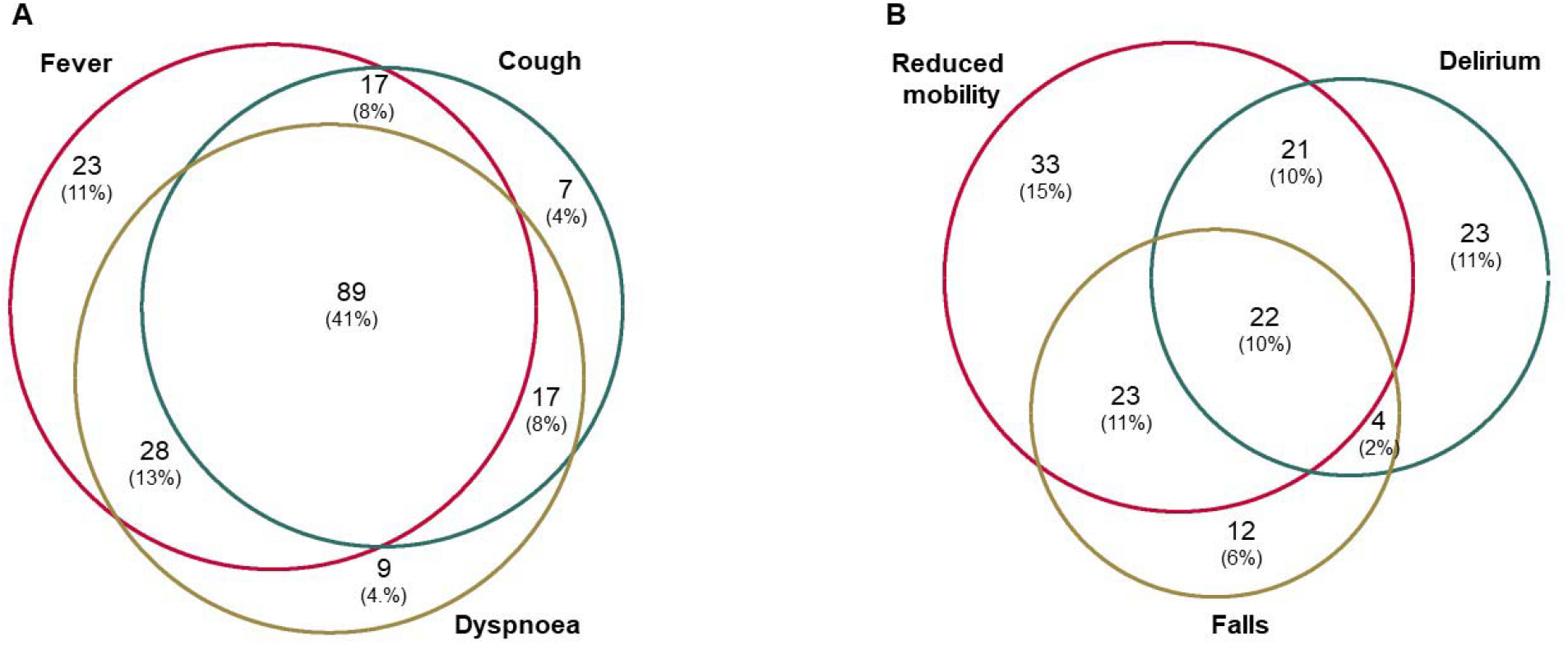
Venn diagram of common presenting clinical symptoms (a) and frailty syndromes (b) in hospitalised older adults with COVID-19.

There was a significant decrease in CRP and neutrophil: lymphocyte ratio with higher CFS scores (Table 2 and Figure 2).

**Table 2.** Distribution of C-reactive protein (CRP) and neutrophil:lymphocyte ratio in mild (Clinical Frailty Scale 1-3), moderate (Clinical Frailty Scale 4-6) and severely frail (Clinical Frailty Scale 7-9) older COVID-19 patients.

|  | **CFS 1-3** | **CFS 4-6** | **CFS 7-9** | **p** |
| --- | --- | --- | --- | --- |
| CRP, mg/L (median, IQR) | 120 (65-250) | 69 (23-169) | 65 (17-119) | <0.01 |
| Neutrophil:lymphocyte<br>(median, IQR) | 7.2 (4.6-17.1) | 6.3 (3.4-9.9) | 5.9 (2.9-9.2) | 0.05 |

**Fig. 2.**
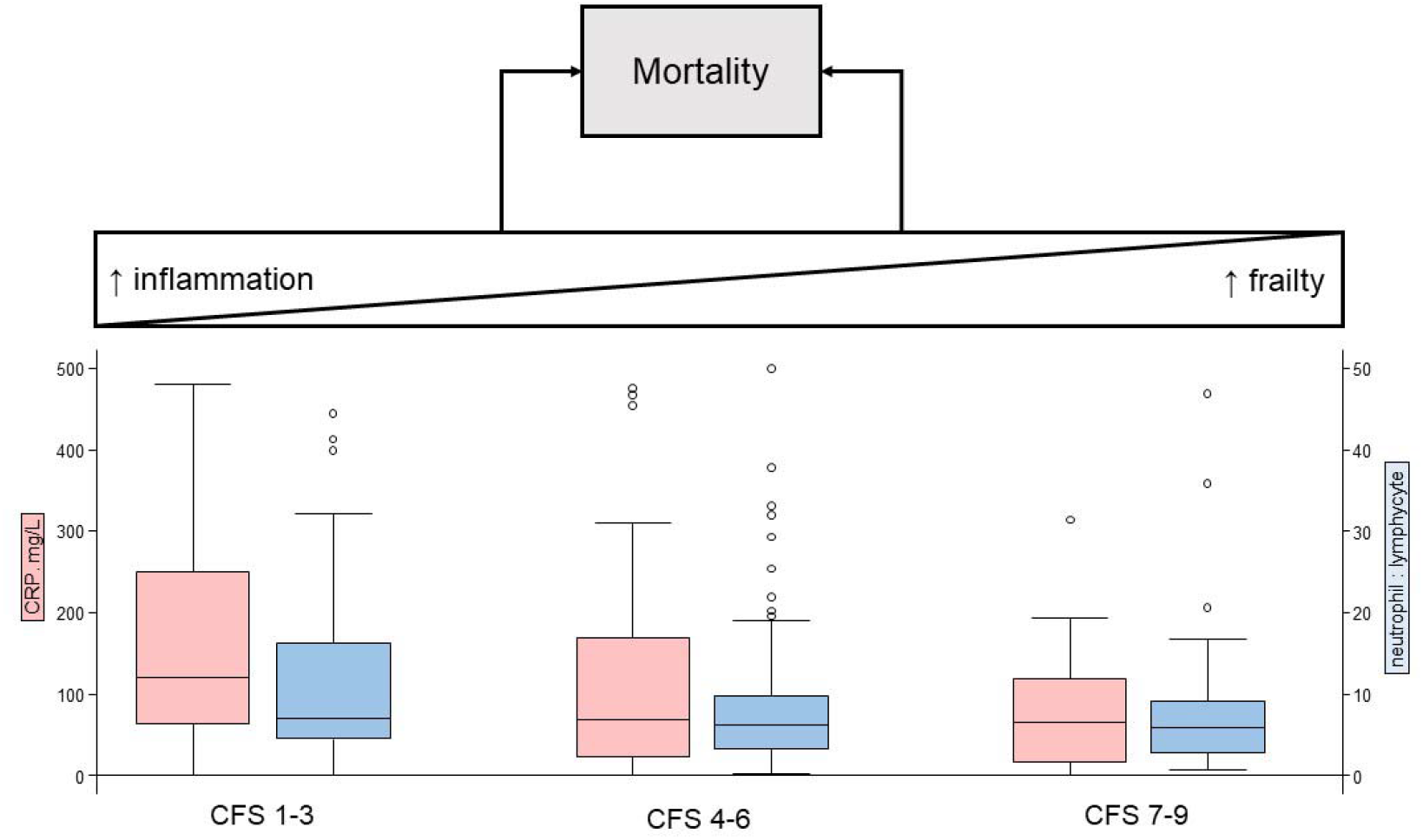
Relationship between immune activity on hospitalisation by degree of frailty and possible divergent routes to mortality.

The contribution of demographic data, clinical presentation and inflammatory markers on admission to mortality was assessed in univariable and multivariable models (Table 3). There was an age-related increase in mortality (HR 1.1, 95% CI 1.0 to 1.1, p<0.01), but neither sex nor frailty was not associated with mortality (Table 3). Of classical COVID-19 symptoms, fever (HR 1.97, 95% CI 1.4 to 3.4, p=0.02) or dyspnoea (multivariable HR 2.0, 95% CI 1.2 to 3.3, p=0.01) at presentation were associated with increased mortality. For frailty syndromes, delirium was associated with mortality (HR 1.9, 95% CI 1.2 to 3.0, p<0.01) (Table 3). With rising CRP or higher neutrophil: lymphocyte ratio there was a corresponding increase in likelihood of death (Table 3).

**Table 3.** Univariable and multivariable analyses estimating the association of presenting features on mortality in older patients with COVID-19.

|  | Univariable models |  |  | Multivariable model |  |  |
| --- | --- | --- | --- | --- | --- | --- |
|  | HR | 95% CI | P | HR | 95% CI | P |
| Age | 1.03 | (1.01-1.06) | 0.02 | 1.06 | (1.03-1.09) | <0.01 |
| Sex | 1.25 | (0.84-1.86) | 0.26 | 1.22 | (0.80-1.87) | 0.36 |
| CFS | 1.02 | (0.93-1.12) | 0.71 |  |  |  |
| Cough | 1.45 | (0.98-2.15) | 0.07 | 1.17 | (0.75-1.84) | 0.49 |
| Fever | 1.75 | (1.09-2.82) | 0.02 | 1.97 | (1.14-3.41) | 0.02 |
| Dyspnoea | 2.14 | (1.37-3.35) | <0.01 | 1.96 | (1.16-3.29) | 0.01 |
| Gastro-intestinal | 0.43 | (0.11-1.74) | 0.24 | 0.45 | (0.11-1.91) | 0.28 |
| Imaging abnormalities | 1.55 | (1.08-2.22) | 0.02 | 1.23 | (0.81-1.86) | 0.33 |
| Falls | 0.91 | (0.59-1.4) | 0.68 | 0.89 | (0.55-1.44) | 0.64 |
| Reduced mobility | 1.1 | (0.76-1.6) | 0.61 | 0.75 | (0.47-1.2) | 0.23 |
| Delirium | 1.28 | (0.87-1.88) | 0.21 | 1.91 | (1.2-3.04) | <0.01 |
| CRP (quartiles) |  |  | <0.01 |  |  | <0.01 |
| 2 | 1.3 | (0.73-2.31) | 0.38 | 1.56 | (0.81-3) | 0.18 |
| 3 | 1.88 | (1.07-3.3) | 0.03 | 2.52 | (1.29-4.94) | <0.01 |
| 4 | 2.37 | (1.38-4.06) | <0.01 | 3.03 | (1.6-5.74) | 0.01 |
| Neutrophil :<br>lymphocytes (quartiles) |  |  | 0.13 |  |  | 0.04 |
| 2 | 0.98 | (0.58-1.68) | 0.96 | 0.86 | (0.47-1.59) | 0.64 |
| 3 | 0.82 | (0.48-1.42) | 0.49 | 0.54 | (0.29-1) | 0.05 |
| 4 | 1.52 | (0.92-2.51) | 0.1 | 1.23 | (0.67-2.25) | 0.5 |

Abbreviations: HR, hazard ratio; CI, confidence interval; CFS, Clinical Frailty Scale; CRP, C-reactive protein

High dependency unit level care and progression to non-invasive ventilation (NIV) or intubation demonstrated higher age-sex-frailty adjusted mortality (HR 3.5, 95% CI, 2.3 to 5.6, p<0.01). In terms of increased rehabilitation needs (evidence of cognitive or physical decline at discharge), only delirium was associated with new or worsening of cognitive impairment in models adjusted for age, sex, dementia and premorbid frailty (OR 44, 95% CI 7.4 to 260). No other admission parameters were associated with cognitive decline or physical decline at discharge, including premorbid dementia.

## Discussion

Here, we describe clinical characteristics and outcomes of older adults admitted to a large London hospital in the first 100 days of the UK COVID-19 outbreak. Frailty syndromes were common presentations, sometimes in the absence of classical COVID-19 symptoms. Some presenting features – fever, dyspnoea, delirium – but not frailty, was associated with increased mortality. However, frailty was associated with a lower degree of inflammation on admission. Taken together, these results quantify the degree to which COVID-19 may present without cardinal symptoms as well as implicate a possible role for age-related changes in immunity (immunesenscence) in mediating the relationship between frailty and mortality.

Our findings should be treated with caution. Our data come from a single site in an urban population in the context of the UK National Health Service (NHS) undergoing restructuring to prepare for the COVID-19 pandemic, limiting generalisation to different health care systems. In particular, the need for ‘further rehabilitation on discharge’ will be specific to the local interface between secondary and community care. Furthermore, as a hospitalised cohort, clinical features and their relation to outcome might vary in population samples. Our measures of immune activity were derived from routinely available laboratory tests on presentation to acute care, rather than direct markers of immunesenescence that might have been available prior to infection. Nonetheless, we had the advantage of specialist geriatrician review of all electronic patient records, which allowed us to ascertain outcomes in near real-time.

Our data indicate that fever and dyspnoea may be important prognostic signs. The clinical significance of delirium has been observed in other COVID-19 cohorts, including from our own hospital [4, 5]. However, the inverse relationship between immune markers on admission and frailty has not previously been noted in COVID-19.

These findings add to the growing descriptive data characterising COVID-19 presentations in older adults [6, 7] and may help inform prognosis at the point of hospital admission. Overt inflammatory activation has been implicated in COVID-19 pathogenesis [8-11] but it is not clear the extent to which this might operate in older adults living with frailty. Frailty and chronic inflammation are linked to immunesenescence and may influence response to infection and subsequent immunity [12-14]. Our finding of lower levels of inflammation in frail patients supports the possibility that background frailty and immunesenescence could constrain the acute inflammation evident in COVID-19 (Figure 2). Whether this accounts for the apparent excess mortality in fitter patients remains speculative [15], though if borne out by further research, has implications for future therapeutic and vaccine strategies.

COVID-19 disproportionately affects older people, warranting a co-ordinated global response. Even in this early stage of understanding the disease, biological complexities that come with ageing are even more apparent in this population. This then becomes an opportunity, indeed a responsibility, for professionals with expertise in clinical and research practice in older people to intensify our efforts on COVID-19.

## Data Availability

On request

## Declarations

### Funding

Daniel Davis is funded through a Wellcome Intermediate Clinical Fellowship (WT107467).

### Conflicts of interest

The authors declare that they have no conflict of interest.

### Ethics approval

These analyses were conducted as part of a service evaluation project and individual consent was not necessary as determined by the NHS Health Research Authority (HRA), the regulatory body for medical research for England, UK. The HRA has the Research Ethics Service as one of its core functions and they determined the project was exempt from the need to obtain approval from an NHS Research Ethics Committee. https://www.hra.nhs.uk/about-us/committees-and-services/res-and-recs/

### Availability of data and material

on request

### Code availability

on request

### Authors’ contributions

PK, AM, TW, NR, IM, BM collected the primary data. DD undertook the statistical analyses and both BW and DD had oversight of the project. PK drafted the first version of the manuscript. All authors contributed to revision and intellectual content of the final submission

## Notes

### Competing Interest Statement

The authors have declared no competing interest.

